# Game-supported cognitive strategy training for slowed information processing speed after acquired brain injury: study protocol for a randomized controlled trial

**DOI:** 10.1101/2022.07.27.22277760

**Authors:** Amy C. Abelmann, Roy P.C. Kessels, Inti A. Brazil, Luciano Fasotti, Dirk Bertens

## Abstract

**Introduction:** Many individuals with acquired brain injury tend to experience problems with slowed information processing speed (IPS). A potentially beneficial and cost-effective supplement for cognitive rehabilitation of impaired IPS may be the implementation of serious gaming that focus on compensatory learning as part of cognitive training. However, most digital platforms that are used during cognitive rehabilitation have been focused on restoration of cognitive function, and limited evidence has been found for the generalization of skills that are trained during cognitive rehabilitation to everyday life.

**Objective:** The aim of this study is to investigate the efficacy of a game-supported cognitive strategy training. The training combines a well-validated time pressure management (TPM) cognitive strategy training targeting slowed IPS with a novel game and a mobile application. The game-supported training focuses on the generalization of strategy-use to untrained tasks in everyday life.

**Methods and analysis:** The study is designed as a randomized controlled trial in which the experimental group (Karman Line Tempo module: an 8-week game-supported cognitive strategy training) will be compared with an active control group (CogniPlus™ training: an 8-week computerized cognitive function training). Data from 60 individuals with acquired brain injury (30 per group, ages between 18 and 70) will be collected at baseline (T0), post-treatment (T1) and at 3-month follow-up (T2). Primary outcome measure is an objective assessment of compensatory strategy-use in an untrained experimental task. Secondary outcome is the attainment of trained and untrained treatment goals assessed by goal attainment scaling (GAS). Pre- and post-training data will be analysed using a 2×2 repeated measure analysis of variance.

**Ethics and dissemination:** This study has been approved by the medical review ethics committee CMO Region Arnhem and Nijmegen (NL74818.091.20) and is registered in the Netherlands Trial Register (NL9437). Research findings will be published in peer-reviewed journals and at conferences.

**Strengths & Limitations:**

- This study is the first to investigate a compensatory approach in serious gaming, combined with a scientifically validated cognitive training, as an intervention for slowed information processing speed in acquired brain injury.
- An adequately powered randomized controlled trial design with block randomization and an active control group to control for confounding effects of computer training in a rehabilitation setting.
- Outcome variables that measure the generalization of the intervention to everyday life, especially to the level of activities and participation, in addition to improvement on neuropsychological tests.
- To balance sufficient power and the feasibility of the study, a sample size of sixty individuals with ABI will be included in the study.
- No 3-month follow-up of the primary outcome measure is included, as only two parallel versions of the Virtual Meeting Task are available.

## Introduction

### BACKGROUND

Individuals with acquired brain injury (ABI) referred for outpatient rehabilitation often experience difficulties with processing and retaining information due to slowed information processing speed (IPS) (1–4). They may experience externally observed slowness (as noticed by others or measured by neuropsychological tests) as well as problems with information processing in everyday activities (5). Consequently, they may experience problems such as feelings of fatigue, lowered mood, and irritability (5). The high frequency and ubiquity of these complaints and their impact on the lives of individuals with acquired brain injury make slowed IPS a prime target for cognitive rehabilitation.

Cognitive rehabilitation in ABI has traditionally focused on the restoration of ‘lost’ cognitive functions, which only has limited effectiveness (6). Compensatory strategy training, however, is more promising as it aims to improve the task performance of individuals within their possibilities and thereby reduce their symptoms by teaching strategies to overcome cognitive deficits (7,8). Compensatory strategy training, as opposed to restorative function training, is currently widely used in the cognitive rehabilitation of people with acquired brain injury (9).

### THE USE OF SERIOUS GAMING IN COGNITIVE REHABILITATION

While compensatory strategy training may be an effective approach for the improvement of performance in daily life tasks, practicing this type of training in face-to-face conditions under guidance of a cognitive trainer is time-consuming and thus costly (10,11). Incorporating elements from serious games (SG) or computerized cognitive training (CCT) in compensatory strategy training may reduce the need for constant supervision as it allows for guided home-practice. We consider the label SG to denote either custom-designed experimental games or commercially designed games with an educative or health-improving approach (12). The label CCT is used to refer to cognitive training programs that integrate game-like features to boost skills or cognitive functioning by repetitively performing demanding tasks over a period of time (13). Most commercially available SG and CCT, such as Lumosity®, HAPPYNeuron©, Rehacom® or CogniPlus™, focus on the restoration of cognitive functions rather than compensation. Such platforms have gained popularity due to their attractive and realistic simulation of everyday situations, with tasks that adapt their difficulty to the level of the individual.

Several studies on the effectiveness of cognitive interventions for acquired brain injury have included SG and CCT. A recent review by Ong and colleagues (12) has shown that the addition of SG to regular cognitive training aimed at ameliorating the impact of for neurological conditions improves rehabilitation outcomes. The implementation of SG in treatment allows for an objective assessment of the individual’s progress (14) and enables more individualized treatment programs tailored to the specific rehabilitation needs (15). Moreover, this implementation leads to better adherence to rehabilitation regimens due to the possibility of guided home practice (16) and improved motivation and engagement (17).

In contrast, studies on CCT in individuals with acquired brain injury have to date shown mixed results with respect to their efficacy and effectiveness in cognitive rehabilitation (18). Most studies found little or no transfer of benefits to non-trained skills or daily functioning (12,19–21), likely due to the focus of the CCT programmes on the restoration of cognitive function rather than compensation for cognitive problems. The beneficial effects of CCT are mostly limited to improvement on trained tasks. In addition, positive effects are short-lasting (22) and many studies have flawed methodology (23–25).

Several recommendations on how to improve investigations in this field were provided by Sigmundsdottir and colleagues (18). They propose the application of a more rigorous methodology, such as the use of more rigid RCT designs, active computer-based control groups, and appropriate activity and participation outcome measures (rather than cognitive test improvement). Combining computer training with the delivery of high dose therapist-assisted interventions may be more effective than isolated CCT or SG, especially when the intervention is a combination of training specific cognitive functions with education on how to apply compensatory strategies to everyday life (26–28).

### GAME-SUPPORTED COGNITIVE STRATEGY TRAINING

The present project aims to investigate the Karman Line Tempo module, a newly developed game-supported cognitive strategy training for brain-injured individuals. The Karman Line Tempo module combines a face-to-face compensatory strategy training for slowed IPS with a custom-designed game set, which consists of an innovative cognitive strategy game (Tempo game) and a mobile application (Tempo tool). An overview of the different components of the Tempo module is presented in Figure 1. Unlike existing SG and CCT, our game set aims to support a shortened version of an already validated face-to-face compensatory strategy training, namely Time Pressure Management (TPM) training (8,29). In TPM training, individuals are taught to identify when they experience time pressure during the execution of a task and to compensate for their slowed information processing speed through strategies to prevent or reduce this time pressure. The individual is taught how to use the TPM strategy under face-to-face guidance of a trained therapist. The addition of the game set to the face-to-face TPM training enables guided home practice: the Tempo game allows for practicing the TPM method in an interactive serious game and the Tempo tool assists during the execution of daily activities by creating a set-by-step plan according to the TPM method.

**Figure 1.**
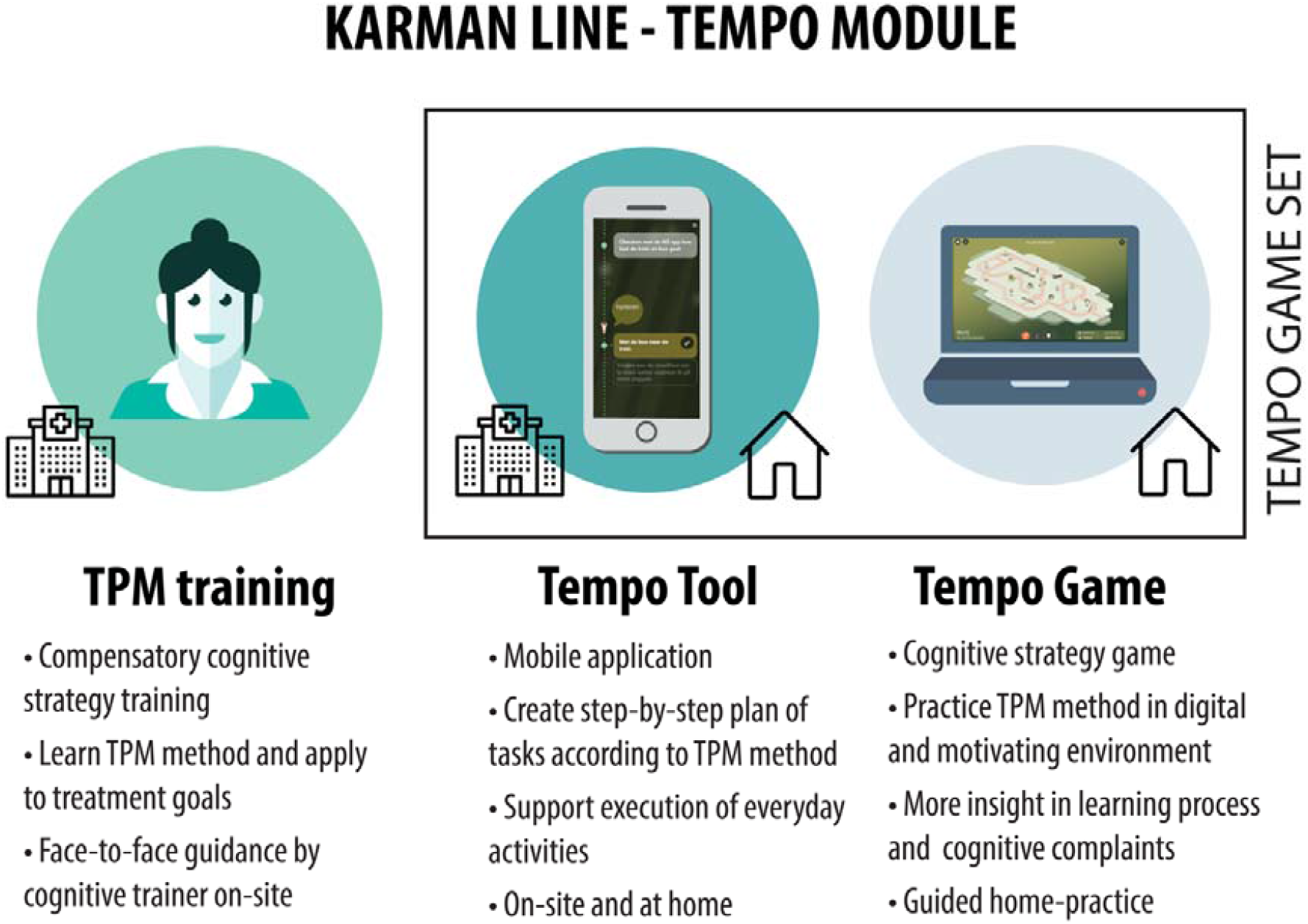
Schematic overview of the Karman Line tempo module

The aim of this protocol paper is to describe the Karman Line Tempo module for individuals with ABI who experience slowed IPS. Moreover, we will describe a randomized controlled trial designed to investigate the efficacy of the Karman Line Tempo module. The main focus of this study is to investigate possible generalization of compensatory strategy-use that was trained during the module to everyday life situations. We hypothesize that the implementation of the Tempo module will lead to a more effective generalization of compensatory strategies to untrained tasks in individuals with ABI relative to an active control group who will receive a computerized cognitive function training. We expect that the Tempo module contributes to improved treatment of slowed IPS in individuals with ABI.

## Methods and analysis

### STUDY POPULATION

The study population consists of individuals with acquired brain injury who are referred for outpatient cognitive rehabilitation. A total of 60 participants will be enrolled in the study. To be eligible to participate, a person must meet all of the following inclusion criteria: A) suffering problems due to slowed IPS (as assessed by a Mental Slowness Questionnaire (MSQ) (30) score ≥ 13) following ABI of nonprogressive nature (i.e. traumatic brain injury, stroke); B) a minimal post-onset time of 3 months after brain injury; C) age between 18 and 70 years; and D) living independently at home. Exclusion criteria of the study are: A) inability to speak/understand the Dutch language; B) a history of severe psychiatric problems; C) neurogenerative disorders; D) substance abuse; E) severe cognitive comorbidity (e.g., dementia), F) inability to look at a computer screen and/or operating a keyboard and/or operating a computer mouse; G) aphasia; and F) visual neglect. Individuals will be recruited from the outpatient clinic and the department of neurorehabilitation of Klimmendaal rehabilitation specialists, Arnhem, the Netherlands. The study has been approved by the medical review ethics committee CMO Region Arnhem and Nijmegen (NL74818.091.20) and is registered at the Netherlands Trial Register (NL9437).

### STUDY DESIGN

The study will be a randomised controlled trial, designed to determine the efficacy of the Karman Line Tempo module: a game-supported strategy training for individuals with ABI aimed at improving compensatory strategy-use for slowed IPS in daily life activities. To achieve this goal, the experimental group (8-week Karman Line Tempo module) will be compared with an active control group (8-week CogniPlus training: a computerized cognitive function training). Participants will be randomly assigned using variable block randomization stratified by gender and age, using Castor Electronic Data Capture (31). Measurements will be administered by a researcher before (T0: baseline) and after the eight-week intervention (T1: post-treatment). The treatment will be given by a trained cognitive trainer or neuropsychologist. A follow-up measurement (T2), consisting of questionnaires, will be obtained three months after treatment. The intervention is single-blinded: investigators were not blinded for group allocation as the nature of the outcome measures ensured a non-biased assessment of performance (e.g., computer obtained measurements). A schematic overview of the study is presented in Figure 2.

**Figure 2.**
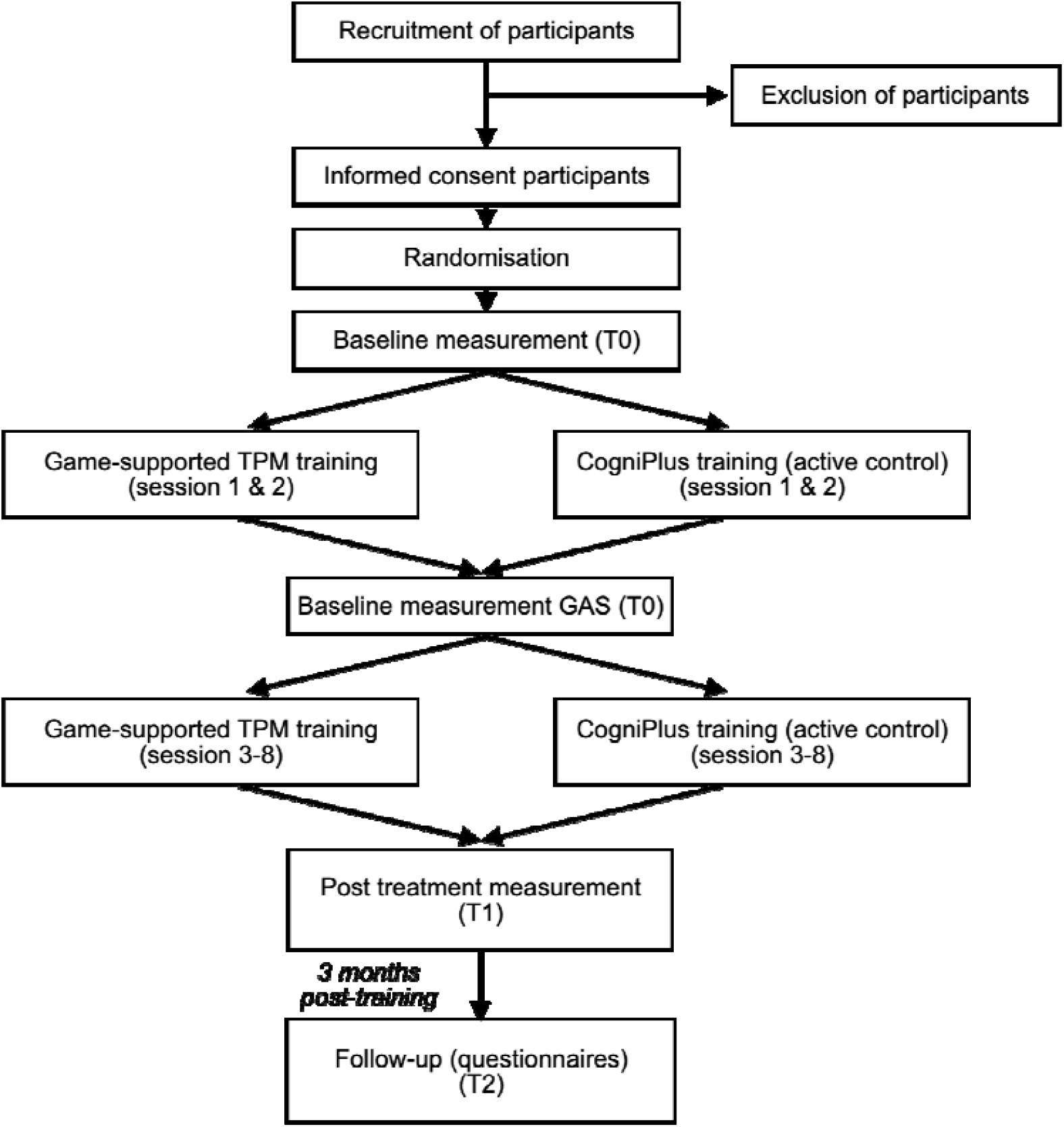
Schematic overview of the study design.

### RECRUITMENT PROCEDURES AND CONSENT

Potential participants are informed about the study and asked to fill in the MSQ prior to a first intake at the rehabilitation centre. Informed consent will be obtained if the individual meets the inclusion criteria and decides to participate after a one-week decision period. Recruitment will take place between December 2021 and July 2023.

### INTERVENTIONS

*Karman Line Tempo module: An experimental game-supported strategy training for slowed IPS* The Karman Line Tempo module is based on the TPM training, a scientifically validated and successfully implemented treatment for individuals who experience slowed IPS (8,29,32). The TPM training focuses on the acquisition of compensatory strategies that allow individuals to recognize and deal with moments of time pressure in everyday situations. The method is based on the general self-instructional idea of “Let me give myself enough time to do a task” (8,29). Trainees are guided by five main questions in the performance of a task while compensating for slowness of information processing. They are taught to 1) distinguish the different steps of a task, to 2) determine which steps are performed under time pressure, to 3) apply strategies to prevent time pressure prior to the task, to 4) deal with imposed time pressure during the execution of the task, and to 5) come up with an emergency plan for unexpected events.

The Karman Line Tempo module combines a shortened version of the face-to-face TPM training with a custom-designed game set and involves eight weekly training sessions (max 60 minutes per session) at the rehabilitation centre (see Figure 1 and Table 1). The game-set consists of an innovative cognitive strategy game (Tempo game) and a mobile application (Tempo tool). The Tempo game is played on a computer and allows for guided home practice of the TPM method in a digital environment. The content of the game follows the pace of the face-to-face TPM training: the individual will only be able to apply the strategies they have learned during the face-to-face training to ensure an optimal learning curve. The Tempo tool is used on a smartphone to create a step-by-step plan, according to the TPM method, and assists in the direct application of compensatory strategies in daily life. The game set is thus not intended as a replacement for the role of the therapist or of the treatment itself, but as an addition to the compensatory cognitive strategy training. The duration of the adapted training in the Tempo module is eight hours as opposed to the ten hours of the original TPM training (29), which is made possible by the engaging nature of the game set and digitally-guided home practice.

**Table 1.** Overview of the Karman Line - Tempo module intervention.

| Description of the Karman Line – Tempo module |  |  |
| --- | --- | --- |
| **Phase 1**<br>Improve insight in slowed IPS,<br>Recognize situations with time pressure | **Session 1**<br>*Information and insight* | Introducing participants to the game-supported TPM training.<br>A brief overview of the purpose of the program and the content of each session.<br>Psychoeducation on slowed IPS and time pressure, secondary problems due to slowed IPS (emotional changes, fatigue, forgetfulness), and the relationship between slowed IPS and impaired performance in daily life.<br>Explain the objective of the TPM game: to practice the time pressure management strategies that are taught during the treatment in a safe and motivating environment.<br>Explain the objective of the TPM tool: to provide support for the execution of daily activities.<br>*No game or tool* |
|  | **Session 2**<br>*Setting up goals* | Defining three personal treatment goals (scored on a Goal Attainment Scale) according to the SMART principle, with support from the practitioner.<br>Two of these goals will be practiced during the treatment, the third one is used to investigate whether training effects transfer to untrained tasks.<br><u>Tempo game (level 2.1 – 2.4)</u>: Introductory. The player learns how to recognize time pressure in four different scenarios. For example, the avatar is lost in a park and has to ask for directions. The patient will learn that listening to the instructions of a passer-by on how to find the nearest exit involves experiencing time pressure. |
| **Phase 2**<br>Acceptance and acquisition of the TPM method | **Session 3**<br>*Introduction TPM method* | Introducing participants to the TPM method aimed at recognizing, preventing and dealing with time pressure.<br>Explain that the patient's IPS does not return to premorbid levels, but that the patient can use the TPM strategy, a 5-step strategy, to compensate for their slowed IPS.<br><u>Tempo game (level 3.1 – 3.4)</u>: Easy difficulty. The player practices with the TPM method by setting up an action plan, and applying different compensatory strategies to prevent and deal with time pressure in different situations. Mini-games include making notes during a conversation, or navigating through a maze while using a timer to impose external time pressure. |
|  | Session 4 & 5<br>*Practice TPM method* | Practicing the TPM method using the two main treatment goals with support from the practitioner.<br>Practicing other situations with time pressure using problem mapping (how do you tackle a task?), listening exercises, roleplay scenarios, and personal situations suggested by the patient.<br><u>Tempo game</u> (level 4.1 – 4.4, and 5.1 – 5.4): Medium difficulty. Level of difficulty increases by increasing the number of task steps, introducing irrelevant steps, introducing the application of an emergency plan and increasing the tempo of the mini-games. For example, the player has to plan a route and navigate through a maze while distractions and difficult decisions are presented in a more rapid tempo. Other mini-games include listening to a conversation between multiple speakers, following instructions to build a rocket ship and preparing a meal.<br><u>Tempo tool</u> The participant uses the tool to set up specific task steps and compensatory strategies for an efficient performance of their two main goals in daily life, and will practice these goals at home and, if possible, during the treatment sessions. |
| **Phase 3**<br>Generalization | Session 6 & 7<br>*Generalization to daily life situations* | Explain that therapists cannot possibly treat all the problems and tasks patients will encounter in their own surroundings. Therefore, generalization of skills and strategies from the rehabilitation to home setting, and from trained to untrained tasks is very important.<br>Discuss and practice situations with time pressure from the patients' daily life.<br>Train various tasks and situations with time pressure under more distracting and difficult conditions.<br><u>Tempo game</u> (level 6.1 – 6.4, and 7.1 – 7.4): Advanced difficulty. Levels become increasingly difficult, player receives less feedback and guidance from the game, tempo of the mini-games are increased, more distractions are introduced and independence of the player's decisions is promoted.<br><u>Tempo tool</u> The tool is applied to new situations in daily life and the patient is encouraged to practice these tasks with help of the tool at home, and if possible, during the treatment sessions. |
|  | Session 8<br>*Evaluation & final session* | Evaluation achievement of personal goals (GAS goals)<br>Evaluation experience of overall treatment<br>Encourage the participant to continue using the TPM method and the Tempo tool in their daily lives. |

To ensure an effective and user-friendly addition to the treatment, the Tempo game set was co-created with patient volunteers with brain injury, therapists and neuropsychological experts. All parties were asked to provide feedback on the tool and the separate game levels throughout the development process. Feedback sessions were organised to explore the experience of the player, to ensure that the game and tool matched the needs and learning objectives of the target group, and improve the usability and design of the game and tool.

The Karman Line Tempo module consists of three main stages. The first stage of the training (sessions 1 and 2) focuses on improving insight into slowed IPS and recognising time pressure. The Tempo game will be used during the first phase to determine a step-by-step plan of an everyday activity and to recognise the steps that invoke time pressure. In the second stage, the TPM method is introduced and three personal treatment goals are set-up (sessions 3–5). The Tempo game will be used to practice the TPM method and apply compensatory strategies in several different situations, such as notetaking during a conversation and navigating in a maze. Three personal treatment goals are set-up using Goal Attainment Scaling (GAS) (33,34). Two of the goals will be trained during the training (using the Tempo tool) and the third goal will remain untrained to investigate whether training effects generalize to untrained tasks. The third stage focuses on the generalisation of the TPM method to functioning in daily life. Again, the Tempo game is used to practice the TPM strategy under more difficult circumstances and the Tempo tool is applied to new daily life activities.

#### Active control group: CogniPlus™ training

The active control group will receive the SPEED module of the CogniPlus™ training program (35), a computerised cognitive training (CCT) programme with a scientific background that aims to train information processing speed (36). No compensatory strategies are taught during this intervention. The CogniPlus™ training program is based on the restorative approach and thus on the assumption that underlying cognitive impairment can be restored, or at least move in the direction of normality, through cognitive exercises (37).

The active control group receives eight weekly 60-minute sessions at the rehabilitation centre (see Table 1). The first two sessions are added prior to the SPEED module and are focused on providing psychoeducation and setting three personal goals according to the GAS scales. From session 3-7 (phase 2), the SPEED module is self-administered on-site while the therapist is present in the room to provide assistance if necessary. During session 8 (phase 3), the individual’s final performance on the training is discussed.

### PRIMARY OUTCOME MEASURE

The primary outcome measure is the Virtual Meeting Task (VMT), an objective assessment of strategy-use in an untrained computerized task that was newly developed specifically for this study (see Figure 3). The VMT examines strategy planning, strategy execution and task execution in a pre-recorded digital setting. The language of the conversations in the task are Dutch, as the task was designed for adults who are living in the Netherlands. The participants will perform parallel, randomized versions of the VMT at baseline level (T0) and at an eight-week follow-up (T1). In the VMT, the participants are instructed to partake in a pre-recorded virtual meeting with four actors and answer questions about the content of the conversation. Participants can use strategies to prevent time pressure (e.g., taking notes) and to manage induced time pressure by selecting strategy buttons that elicited a response (e.g., asking for repetition). After each scene, the participants answer questions on the content of the conversation. Performance in the VMT will be scored on the following aspects: strategy planning (number of correctly chosen strategies that can prevent time pressure), strategy execution (number of correctly chosen strategies that help manage time pressure) and task execution (number of correct reproductions of information points). After completing the task, the participants perform a self-rating questionnaire which measures personal insight on the subjective experience of perceived task performance and cognitive state. Construct validity, test-retest reliability, parallel form reliability and ecological validity of the experimental task will be investigated in a study that runs simultaneously with the study described in this protocol.

**Figure 3.**
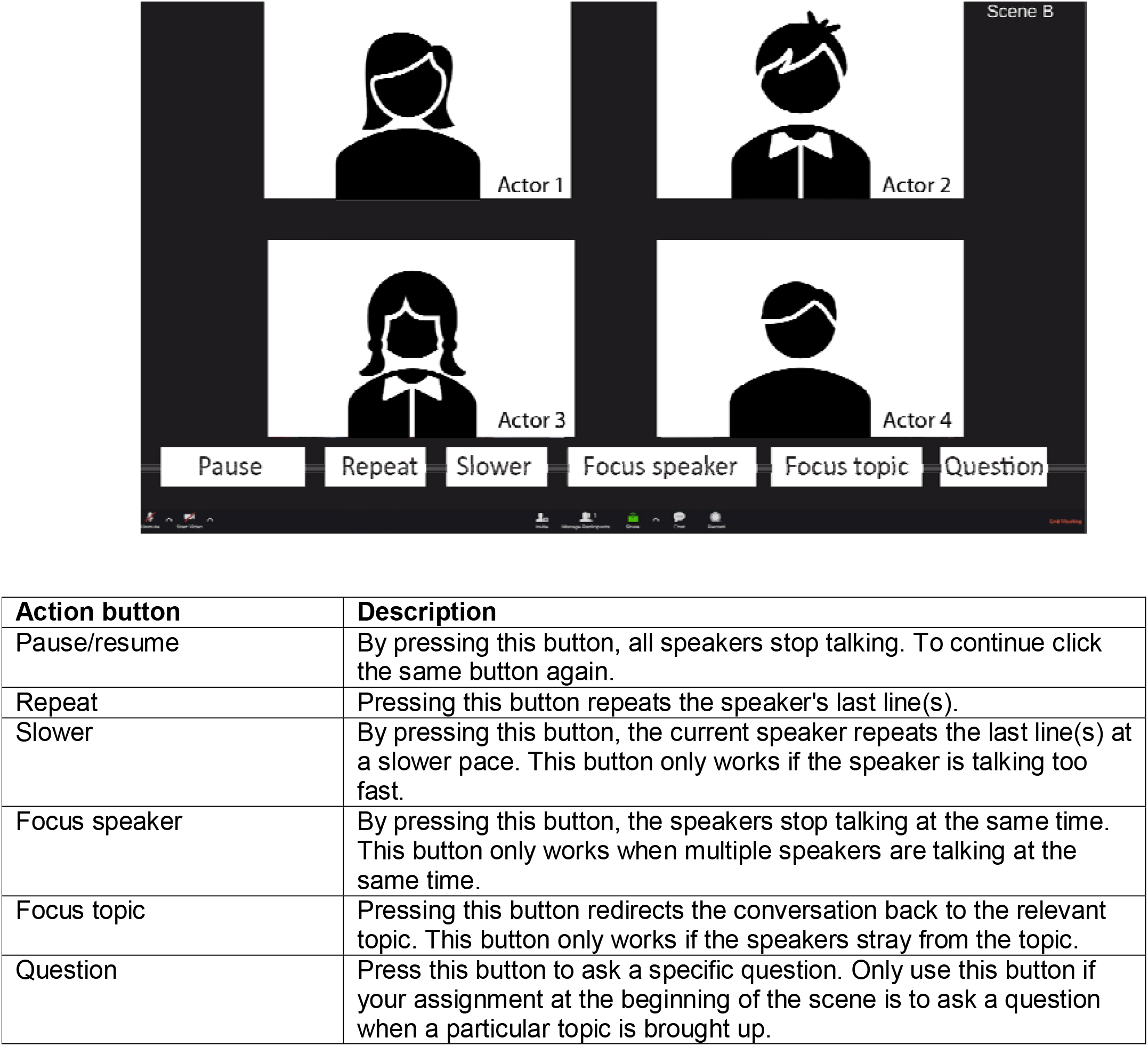
Overview and task outcomes of the Virtual Meeting Task for objective assessment of compensatory strategy-use.

### SECONDARY OUTCOME MEASURE

The secondary outcome measure is a standardised Goal Attainment Scaling score (33,34). GAS is an individualised method used in brain injury rehabilitation to evaluate to what extent individual treatment goals are achieved. The achievement of each goal will be measured on a 5-point scale (38,39). In fact, each individual has their specific treatment goals, but this is scored in a standardised way to allow statistical analysis. For the present study, three treatment goals will be identified that relate to situations in which the individual experiences problems due to slowed IPS. Acceptable treatment goals can be subdivided into multiple steps and are defined according to the SMART principle (Specific, Attainable, Reasonable, Timely) (34,40).

### ADDITIONAL STUDY PARAMETERS

Demographic measures include age, sex, education level, type of brain injury, and time after injury. Other study parameters include self-rating questionnaires measuring compensatory strategy-use in everyday life (Time Pressure Questionnaire (32)), participation in daily life (Utrecht Scale for Evaluation of Rehabilitation-Participation (41); USER-P), subjective motivation (Motivation for Traumatic Brain Injury Rehabilitation Questionnaire (42,43); MOT-Q), subjective fatigue (Dutch Multifactor Fatigue Scale (44); DMFS), and insight and reported severity of mental slowness (MSQ (30)). A selection of questionnaires (Time Pressure Questionnaire, MSQ, USER-P, DMFS) will be administered at 3-month follow-up to assess the long-term effects of the training.

A neuropsychological test battery will be administered to assess information processing speed and other aspects of cognitive functioning at baseline. For this purpose, subtests of the Test for Attentional Performance (TAP 2.3.1) (45) will be administered. Specifically, the Alertness subtest to assess general processing speed, the Sustained Attention subtest to assess the longer-term maintenance of attention, the Working Memory subtest to assess the ability to continuously update the content of working memory, and the Go/No-go subtest to assess cognitive control. The Attention Network Test (ANT) (46) is used to assess the severity of slowed IPS. The Brixton Spatial Anticipation Test (47) will be administered to assess set-shifting and perseveration. The Dutch version of the National Adult Reading Test (NART) (48,49) is included to estimate premorbid IQ. A selection of neuropsychological tests will be repeated post-treatment to control for potential nonspecific recovery. An overview of the measurements is presented in Table 3.

**Table 2.** Overview of the CogniPlus™ training intervention.

| **CogniPlus™ training** |  |  |
| --- | --- | --- |
| **Phase 1**<br>Improve insight in slowed IPS<br>Introduction CogniPlus™ training program | **Session 1**<br>*Information and insight* | Introducing participants to the CogniPlus training program.<br>A brief overview of the purpose of the program and the content of each session.<br>Psychoeducation on slowed IPS and time pressure, secondary problems due to slowed IPS (emotional changes, fatigue, forgetfulness), and the relationship between slowed IPS and impaired performance in daily life.<br>Explain the objective of the SPEED module of the CogniPlus training program, that is, to improve problems due to a slowed IPS by practicing computer tasks that are aimed at improving the speed of information processing. |
|  | **Session 2**<br>*Setting up goals* | Setting up three personal treatment goals (scored on Goal Attainment Scale) according to the SMART principle, with support from the practitioner.<br>Two of these goals will be practiced during the treatment, the third one is used to investigate whether training effects transfer to untrained tasks. |
| **Phase 2**<br>CogniPlus™ training program | **Session 3 – 7**<br>*Training with SPEED module* | The patient will spend one hour per week practicing different tasks of the SPEED module using three scenarios.<br>The tasks from the scenario's consist of ordering objects in ascending order, sorting objects on shelves, and avoiding obstacles on a track. All of these scenarios impose time pressure and the patient is encouraged to increase their reaction time and speed of actions.<br>The patient can switch between scenarios in a self-directed manner.<br>The practitioner will be present in the room to help the patient if they have any questions and to track their progress.<br>After the patient is finished with practicing the tasks, the practitioner will reflect together with the patient on the performance based on the results of the training session. |
| **Phase 3**<br>Final results SPEED module | **Session 8**<br>*Evaluation & final session* | The patient plays all scenarios from the SPEED module one last time.<br>Final feedback is given on the participant's progress throughout the training.<br>Evaluation achievement of personal goals (GAS goals).<br>Evaluation experience of overall treatment. |

**Table 3.** Inclusion and outcome measures.

| Type | Measurement | Inclusion | Base-line | Post-treatment | Follow-up |
| --- | --- | --- | --- | --- | --- |
| **Primary outcome measure** | Online Meeting Task |  | x | x |  |
| **Secondary outcome measure** | Treatment goals (GAS) |  | x | x |  |
| **Neuropsychological tests** | *Test Battery for Attentional performance (TAP)*<br>- Alertness<br>- Working memory<br>- Sustained attention<br>- Go/No-go task |  | x<br>x<br>x<br>x<br>x | x<br>x |  |
|  | Brixton test |  | x |  |  |
|  | Attention Network Test (ANT) |  | x | x |  |
|  | National Adult Reading test |  | x |  |  |
| **Questionnaires** | Mental Slowness Questionnaire (MSQ) | x | x | x | x |
|  | Utrecht Scale for Evaluation of Rehabilitation-Participation (USER-P) |  | x | x | x |
|  | Strategy-use in individuals with slowed IPS (Questionnaire “Omgaan met tijdsdruk”) |  | x | x | x |
|  | Dutch Multifactor Fatigue Scale (DMFS) |  | x | x | x |
|  | Motivation for Traumatic Brain Injury Rehabilitation Questionnaire (MOTQ) |  | x | x |  |

Moreover, data will be obtained from the Tempo game to evaluate the performance of the individuals in the experimental group in the game, such as reaction times and the number of correct responses. Based on this data, variables are created to estimate strategy-use, understanding of the game, attention, fatigue, perseveration and motivation. Predictions are made by a general probabilistic scaling method underlying the game that can estimate variables based on their posterior probabilities (50). The validity of this diagnostic network will be investigated in another study that runs simultaneously with the study described in this protocol.

### DATA MANAGEMENT

Study data will be collected and managed using Castor Electronic Data Capture (EDC) (31) hosted at the Donders Institute for Brain, Cognition and Behaviour, Nijmegen, the Netherlands. Castor EDC is a secure, web-based software platform designed to support data capture for research studies.

Participants will be identified using a study-specific identification code. One researcher will keep a separate participant identification code list in the protected project folder-based at the sponsor’s site (rehabilitation centre Klimmendaal, Arnhem, the Netherlands) that matches the study-specific identifying codes with the participants’ names. Paper documents are kept in a locked safe at the sponsor’s site.

### PATIENT INVOLVEMENT

We involved individuals with ABI and members of the public in several stages of the development of the game-supported treatment. To ensure an effective and user-friendly addition to the treatment, the Tempo game set was co-created with patient volunteers with brain injury, therapists and neuropsychological experts. All parties were asked to provide feedback on the tool and the separate game levels throughout the development process. Feedback sessions were organised to explore the experience of the patient, to ensure that the game and tool matched the needs and learning objectives of the target group, and improve the usability and design of the game and tool.

Together with rehabilitation experts we carefully assessed the implementation of the trial interventions to minimize the burden on the individuals with ABI. Our intention is to inform the participants of the main results of the study and involve individuals with brain injury in the discussion of appropriate methods to disseminate the research outcomes.

### SAMPLE SIZE

A power calculation for the primary outcome measure was based on the results of a prior study that used a similar compensatory strategy task (the Information Intake task), study design, and research population (51). We based our expected standardized effect size of at least 0.66 on the effect size obtained in the study above. We calculated that a selected sample of 60 participants (30 per group) will be sufficient to reach a power of .8 (α=.05).

### ANALYSIS OF PRIMARY OUTCOME MEASURE

To evaluate the efficacy of the game-supported treatment compared to the active control group pre- and post-training data obtained in the Virtual Meeting Task will be analysed using a 2×2 repeated measure analysis of variance (ANOVA) with training Group (experimental group and active control group) as between-subject factor and Measurement (2 levels: pre-treatment, post-treatment) as within-subject factor. Three aspects will be measured by the experimental task (strategy planning, strategy execution and task execution). The individual aspects of the task will be transformed into standardized scores and grouped into a single compound measure score as a primary outcome measure.

### ANALYSIS OF SECONDARY OUTCOME MEASURE

As a secondary outcome measure, pre-and post-training GAS scores of the three (un)trained goals will be analysed using a 2×2 repeated measure analysis of variance (ANOVA) with training Group (game-supported treatment and active control group) as between-subject factor and Measurement (2 levels: pre-and post-treatment) as within-subject factor.

### ANALYSIS OF ADDITIONAL STUDY PARAMETERS

Correlations will be computed between moderator variables and the treatment effects (difference score post-minus pre-treatment). Regression analysis will be performed to investigate the relation between the measured outcomes of the game (error rates, reaction times) and baseline measures of neuropsychological assessment and questionnaires.

Path analyses will be performed to assess the effectiveness of the Bayesian network that underlies the Tempo game in creating predictions of cognitive values obtained in the game (i.e., decrease in attention). Predictive models will be evaluated to examine the relationships between the variables obtained in the Tempo game and the outcomes on neuropsychological tests. Four predictive models will be tested. For the first model, the game variable that was obtained for strategy-use was used as a predictor of the strategy execution, task execution and strategy planning score of the Virtual Meeting Task. The second model uses the game variable for attention as a predictor of the Alertness subtest and the Sustained Attention subtest of the TAP. The third model makes use of the game variable for fatigue as a predictor of the course of RT of the Alertness subtest of the TAP and the outcome of the Dutch Multifactor Fatigue Scale questionnaire. For the fourth model, we used the game variable for perseverance as a predictor for perseverative errors on the Brixton Spatial Anticipation test.

### ETHICS AND DISSEMINATION

This study has received ethical approval from CMO Region Arnhem-Nijmegen medical research ethics committee (NL74818.091.20) and is registered in the Netherlands Trial Register (NL9437). Data are handled confidentially and anonymously. Where it is necessary to be able to trace data to an individual participant, a subject identification code list can be used to link the data to the individual. The principal investigator will safeguard the key to the code. The data will be stored for 15 years. The handling of personal data is in compliance with the European General Data Protection Regulation (GDPR). The study will be conducted according to the principles of the Declaration of Helsinki (64th WMA General Assembly, Fortaleza, Brazil, October 2013) and in accordance with the Medical Research Involving Human Subjects Act (WMO). All participants in the active control group are offered the standard TPM training at Klimmendaal rehabilitation centre after completion of the study.

Dissemination among the academic community and general public will involve publications in international, peer-reviewed journals, presentations at leading national and international conferences, media appearances in (local) newspapers, on social media, online blogs and public events. Publication will not be limited to positive findings.

## Discussion

This randomized controlled trial aims to investigate the efficacy of the Karman Line Tempo module, a game-supported cognitive strategy training for individuals with ABI who experience slowed IPS. The goal of this study is to 1) examine the generalization of strategy-use as assessed in an untrained experimental task, and 2) investigate individual subjective experience of achievement of trained and untrained treatment goals as measured by standardized GAS.

The main strength of this study is the novelty of the compensatory approach in the Tempo module. This study is the first to investigate a game-supported cognitive strategy training that implements novel custom-designed digital platforms with a compensatory approach for ABI rehabilitation. Existing CCT and SG interventions mainly focus on improvement of cognitive function and previous research has shown limited generalization of skills to everyday life activities (12,19– 22,37). The Tempo module overcomes these limitations by combining both face-to-face compensatory cognitive training sessions with a custom-made digital game (Tempo game) and a mobile application (Tempo tool) that enables home-guided practice. All components of the Tempo module focus on teaching individuals compensation strategies for slowed IPS and guiding them in applying these strategies in everyday life, providing them with a robust basis for long-term improvement.

Another strength is the experimental design of the study, which incorporated the recommendations formulated in the review by Sigmundsdottir et al. (18). The authors criticized the insufficient methodological quality of previous research that used CCT as an intervention in ABI. Previous studies showed a weak methodical design and a lack of active control groups. We apply an adequately powered RCT design and implement the CogniPlus™ training program as active computer-based control group. We also include outcome variables that measure the generalization of the intervention, especially to the level of activities and participation, in addition to improvement on neuropsychological tests.

In summary, the aim of this study is to examine the efficacy of the Karman Line Tempo module for individuals with ABI who experience slowed IPS, focusing on generalization of compensatory strategy-use in everyday life situations. This study could contribute to upgrade the treatment of slowed IPS in cognitive rehabilitation and, more generally, improve our insight into the implementation of digital platforms in cognitive training.

## Data Availability

This manuscript describes a research protocol paper. All data produced in the future will be available online in the Donders Repository (https://data.donders.ru.nl/).

## Acknowledgements

The authors would like to thank Yellow Riders, GameArchitect and Big4Data for designing and developing the Karman Line – Tempo game set. The authors would like to thank Thijs Doornbos, who programmed the Virtual Meeting Task in collaboration with ACA.

## Author Contributions

ACA developed the intervention, designed the study and wrote the first draft of the article. DB, RK and LF reviewed and extended the draft. IB provided support in the set-up of the analysis of the study. All authors critically reviewed the content and approved the final manuscript.

## Patient consent

Not required.

## Ethics approval

Medical review ethics committee CMO Region Arnhem and Nijmegen (NL74818.091.20).

